# Early, low-dose hydrocortisone and near-term brain connectivity in extremely preterm infants

**DOI:** 10.1101/2022.11.25.22282693

**Authors:** Sarah E. Dubner, Lucy Rickerich, Lisa Bruckert, Rocío Velasco Poblaciones, Dawson Sproul, Melissa Scala, Heidi M. Feldman, Katherine E. Travis

**Author notes:** Corresponding Author: Katherine E. Travis, Developmental-Behavioral Pediatrics, Stanford University School of Medicine, 3145 Porter Drive, MC 5395, Palo Alto, CA 94304. **Category of Study:** Clinical.

## Abstract

**Background:** Postnatal steroids are used to prevent bronchopulmonary dysplasia in extremely preterm infants but may have adverse effects on brain development. This study assessed variation in connectivity metrics of major white matter pathways in the cerebrum and cerebellum at near-term gestational age among infants who did or did not receive a standardized regimen of hydrocortisone during the first 10 days of life.

**Methods:** Retrospective cohort study. Participants: Infants born < 28 weeks, divided into 2 groups: Protocol group (n=33) received at least 50% of and not more than 150% of an intended standard dose of 0.5mg/kg hydrocortisone twice daily for 7 days, then 0.5 mg/kg per day for 3 days; versus Non-Protocol group (n=22), that did not receive protocol hydrocortisone or completed <50% of the protocol dose. We assessed group differences in near-term diffusion MRI mean fractional anisotropy (FA) and mean diffusivity (MD) across the corticospinal tract, inferior longitudinal fasciculus, corpus callosum and superior cerebellar peduncle.

**Results:** Groups were comparable in terms of gestational age, post-menstrual age at scan, medical complications, bronchopulmonary dysplasia, and necrotizing enterocolitis. No significant large-effect group differences were identified in mean FA or MD in any cerebral or cerebellar tract between the two groups.

**Conclusion(s):** Low dose, early, postnatal hydrocortisone was not associated with significant differences in white matter tract microstructure at near term gestational age.

**Impact:**

- This study compared brain microstructural connectivity as a primary outcome among extremely preterm infants who did or did not receive early postnatal hydrocortisone.
- Low dose hydrocortisone in the first 10 days of life was not associated with significant differences in white matter microstructure in major cerebral and cerebellar pathways
- Hydrocortisone did not have a significant effect on early brain white matter circuits.

## INTRODUCTION

Bronchopulmonary dysplasia (BPD) is a severe complication of prematurity, characterized by chronic pulmonary inflammation, systemic inflammatory response, and associated local inflammation in the brain.^1–3^ Preterm (PT) infants who go on to develop BPD are proposed to have early hypo-responsiveness of the hypothalamic-pituitary-adrenal axis and are unable to secrete sufficient cortisol in response to inflammatory triggers at the time of PT birth.^4^ Preterm children who develop BPD are at heightened risk for experiencing further health complications and neurodevelopmental delays.^5^

Early hydrocortisone administration, soon after birth, as described in the PREMILOC study, is an intervention aimed at preventing BPD by addressing this relative adrenal insufficiency.^6^ In the PREMILOC study, no differences were reported in cerebral volumes of treated infants at near-term, and neurodevelopment at 2 years was comparable overall and improved for those born at 24-25 weeks, suggesting that the treatment is not deleterious.^7–9^ However, the effect of IV hydrocortisone on early white matter brain development, which has important links to neurodevelopment^10–12^ remains unknown.

Potent glucocorticoids, such as dexamethasone, have been shown to have deleterious effects on brain development.^13–15^ Hydrocortisone is 25 times less potent than dexamethasone and mechanistically mimics naturally-occurring cortisol. Like other glucocorticoids, hydrocortisone may delay maturation of myelin producing precursor cells, pre-oligodendrocytes, and confer an adverse effect on white matter development. However, hydrocortisone’s reduction of systemic inflammation may result in less damage to pre-oligodendrocytes than the infant would have otherwise experienced without treatment. How this balance of potential positive and negative effects plays out in the developing infant brain remains unclear. Inflammation-induced brain white matter microstructural alterations are detectable by term-equivalent age using advanced diffusion MRI (dMRI) neuroimaging techniques^16^ even in the apparent absence of gross injury on clinical MRI.^3^ The sensitivity of dMRI to white matter microstructural alterations from inflammation could inform clinical care of the possible effects of hydrocortisone on brain development.

No studies to date have investigated the effects of early postnatal hydrocortisone on the microstructure of white matter brain tissue in preterm infants. The objective of this investigation was to assess differences in white matter microstructural metrics, as indexed by diffusion MRI metrics (dMRI) fractional anisotropy (FA) and mean diffusivity (MD), of major white matter circuits at near-term equivalent age between two groups of infants born extremely preterm (< 28 weeks gestational age) who received either early, low-dose hydrocortisone treatment per the PREMILOC protocol or did not receive protocol steroids. Given the lower potency of hydrocortisone and its beneficial effects on the infant’s inflammatory response, we hypothesized that protocol-treated infants would have higher FA and lower MD, suggestive of healthy white matter development, in the major cerebral and cerebellar white matter tracts compared to non-protocol infants. Given that our study is a novel use of dMRI for assessing outcomes of neonatal medication use, we were interested in identifying group differences in tract measures with large effect sizes (Cohen’s d >0.8), because such differences would be likely to have clinical interpretability.

## METHODS

### Participants

Children <28 weeks gestation at birth who were born at the Lucile Packard Children’s Hospital (LPCH) Stanford NICU between May 1, 2016 and April 30, 2021 were eligible for this retrospective study. Per LPCH NICU clinical standards of care, all extremely preterm infants undergo routine dMRI imaging near hospital discharge. MRIs were obtained when infants were stable in an open crib, requiring no more than low flow supplemental oxygen for respiratory support, and >34 weeks PMA. Sedation was not used, and scanning was performed during natural sleep. Infants were fed and swaddled in warm blankets and wore adhesive foam noise attenuators to reduce the sound from the scanner. N=100 infants < 28 weeks gestation underwent routine dMRI imaging near hospital discharge. For this analysis, n=45 infants were excluded for several reasons: evidence of genetic disorders or congenital brain anomalies (n=2), received steroids outside the protocol time frame (n=11), MRI scan completed more than one month post-due date (n=1), T1-weighted scan issues (n = 1), incomplete diffusion scans (n = 10), diffusion scan quality issues (e.g., challenges with image processing due to low gray-white matter contrast n = 20). The final sample was 55 participants, shown in **Figure 1**.

**Figure 1.**
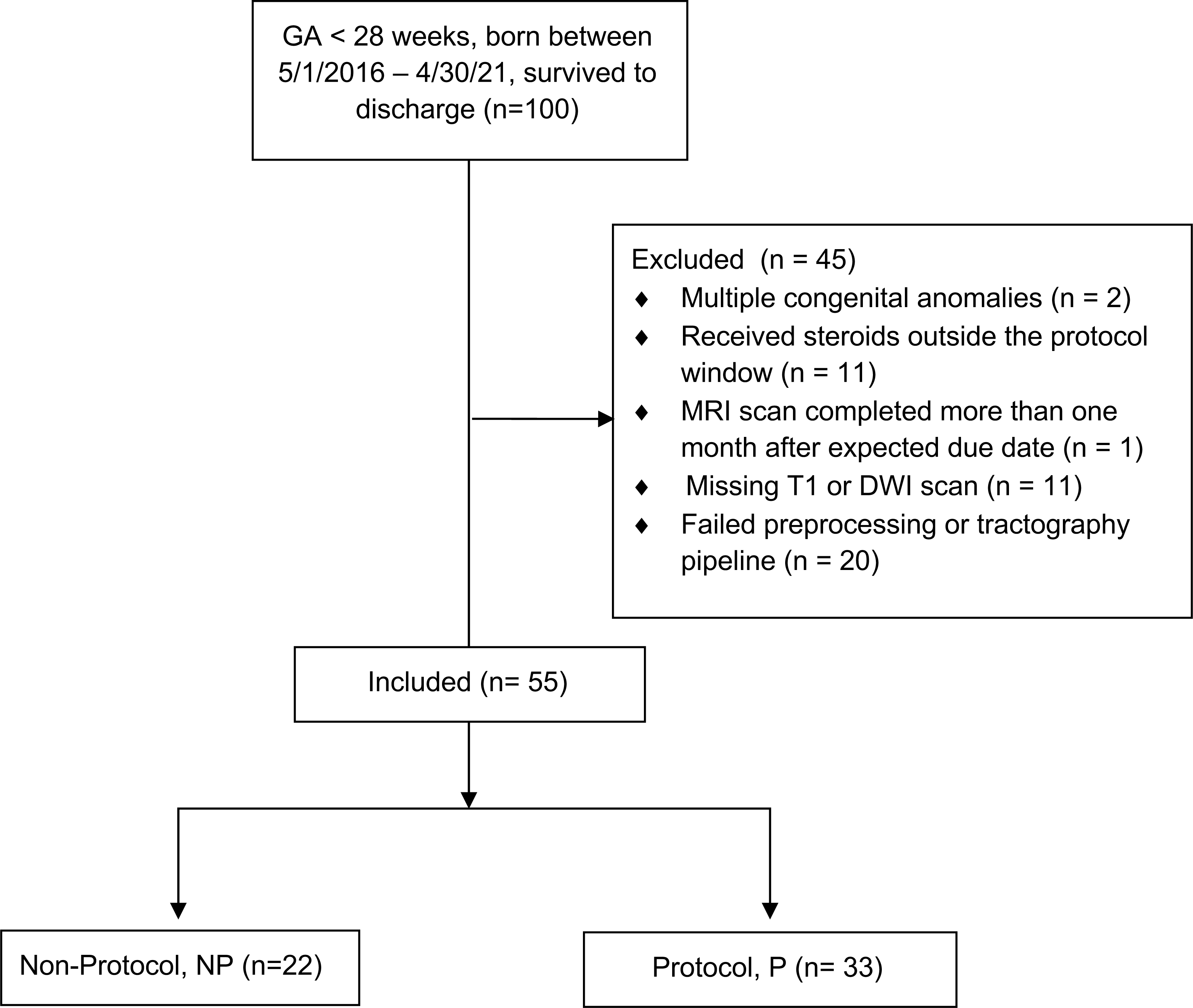
Consort diagram of eligible and included infants.

Infants were categorized by steroid exposure into two groups: Protocol (P) or Non-Protocol (NP). From July 1, 2017 – December 31, 2019 infants born at < 28 weeks gestation were typically begun on early postnatal hydrocortisone following the PREMILOC protocol described by Baud et. al.^6^: starting on day of life 1, 1 mg/kg of hydrocortisone hemisuccinate per day divided into two doses per day for 7 days, followed by one dose of 0.5 mg/kg per day for 3 days. Within the protocol timeframe, discontinuation of or additional hydrocortisone administration was at the discretion of the treating clinician. None of the participants received more potent steroids, such as dexamethasone. The P group (n=33) received greater than 50% of the hydrocortisone protocol; 24 infants received the full hydrocortisone protocol; 5 additional infants who were started on the protocol received 50% to 99% of the total protocol dose due to doses held for hypertension (n=2), hyperglycemia (n=2), or missed doses (n=1); 4 additional infants received 101% to 150% of the total protocol dose, receiving the PREMILOC protocol plus additional hydrocortisone during the first 10 days of life. The NP group (n=22) qualified for the study and received less than 50% of the total protocol steroid dose; 16 infants had no steroid exposure; 6 additional infants began the protocol but did not complete it due to hyperglycemia (n=4), hypertension (n=1), or improving respiratory status (n=1). Clinical variables were obtained via chart review. This retrospective study was approved by the Stanford University Institutional Review Board #IRB-44480.

### MRI Acquisition

The neuroimaging data collected as part of the routine scanner sequence for each infant that were used in this study include high-resolution T1-weighted anatomical MRIs and diffusion MRI data. MRI scans were acquired using a GE Discovery MR750 3.0T scanner equipped with either an 8-channel HD head coil (n = 40) or a 32-channel HD head coil (n = 15) (General Electric Healthcare, Little Chalfont, UK). Across the full sample, no significant difference in FA and MD values were seen when comparing between head coil types in this cohort.

The MRI protocol included a 60-direction diffusion MRI scans with b-values (1,500sec/mm^2^) collected with a multi-slice echoplanar imaging (EPI) protocol for rapid image acquisition (∼3 min each, 6 min total) with two - six volumes at b=0. Diffusion MRI data were collected at 2.0mm^3^ spatial resolution (2.0mm x 2.0mm x 2.0mm isotropic voxels). High-resolution T1-weighted scans were collected at ∼1mm^3^ spatial resolution. T1-weighted images were used as participant-specific anatomical references throughout the processing of diffusion MRI data.

### Diffusion MRI analyses

MRI data were managed and analyzed using a neuroinformatics platform (Flywheel.io). Diffusion MRI data preprocessing and tractography were performed using procedures implemented in Reproducible Tract Profiles (RTP) (https://github.com/vistalab/RTP-pipeline).^17,18^ RTP consists of three main steps: (i) Structural processing and Region of Interest (ROI) creation (ii) diffusion MRI preprocessing (RTP-preproc) and (iii) whole-brain tractography and tract segmentation (RTP-Pipeline). Detailed information for processing steps were described in detail by Lerma-Usabiaga et al.^17^ and Liu et al.^18^ We modified RTP to perform segmentation of the T1w image using Infant Freesurfer^19^ (https://surfer.nmr.mgh.harvard.edu/fswiki/infantFS) to accommodate for the immature neonate brain. We substituted a neonatal template (Edinburgh Neonatal Atlas, ENA33)^20^ with ROIs for tract identification and segmentation.

We performed the following diffusion image preprocessing steps (RTP-preproc): (i) data denoising with principal component analysis^21,22^ and Gibbs ringing correction,^23^ (ii) eddy current and motion correction,^24^ (iii) and anatomical alignment of diffusion data to the average of the non-diffusion-weighted volumes, which were registered to the infant’s high-resolution ac-pc aligned anatomical image using rigid body transformation. Diffusion preprocessing, modeling and tractography within RTP is largely based on software from FSL (https://fsl.fmrib.ox.ac.uk/fsl/fslwiki), MRTrix3 (https://www.mrtrix.org/) and Automated Fiber Quantification^25^ (AFQ, https://github.com/yeatmanlab/AFQ). A full list of software dependencies can be found here (https://github.com/vistalab/RTP-pipeline/wiki).

The preprocessed diffusion MRI data output from RTP-preproc served as the input for diffusion metrics modeling, whole-brain tractography and tract segmentation in RTP-pipeline. White matter metrics were calculated based on the diffusion tensor model. We then used the constrained spherical deconvolution model (CSD),^26^ with eight spherical harmonics (lmax = 8) to calculate fiber orientation distributions (FOD) for each voxel. CSD tractography and FA masks were adjusted for the immaturity of the neonatal brain to have FA mask thresholds of 0.15 with FOD of 0.08. The CSD FODs were used for diffusion MRI tractography, which consisted of (i) Ensemble Tractography^27^ to estimate the whole-brain white matter connectome. MRtrix3 was used to generate three candidate connectomes that varied in their minimum angle parameters (50°, 30°, 10°).^28^ For each candidate connectome, a probabilistic tracking algorithm (iFOD2) was used with a step size of 1 mm, a minimum length of 10 mm, a maximum length of 200 mm, and an FOD stopping criterion of 0.04. (ii) Spherical-deconvolution Informed Filtering of Tractograms (SIFT) to improve the quantitative nature of the ensemble connectome by filtering the data such that the streamline densities match the FOD lobe integral. The resulting ensemble connectome retained 500,000 streamlines. (iii) Concatenation of the three candidate connectomes into one ensemble connectome. (iv) AFQ^25^ to segment and refine the resulting whole-brain connectome of each child into the tracts of interest: the left and right corticospinal tract (CST), left and right inferior longitudinal fasciculus (ILF), left and right superior cerebellar peduncle (SCP), and the anterior, middle, and posterior segments of the corpus callosum (Anterior Frontal, Motor, Occipital).^29,30^ **Figure 2** shows the ROIs and resulting tracts on a T1 image from a 26 3/7 week infant imaged at 38 2/7 weeks PMA. Mean tract FA and MD were calculated for the core tract area between defining ROIs.

**Figure 2.**
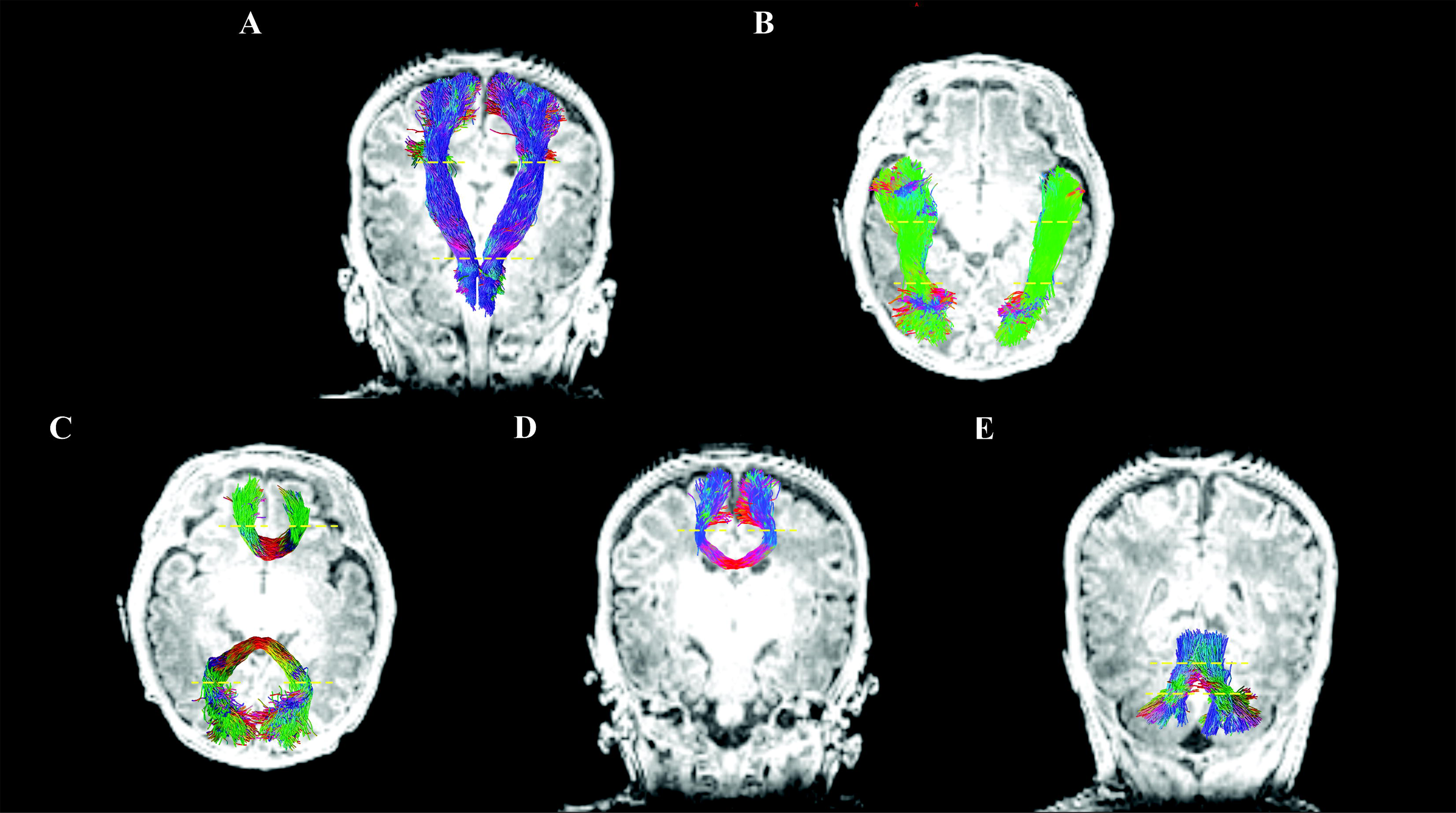
White Matter Tractography. A) Left and right Corticospinal Tract, B) Left and right Inferior Longitudinal Fasciculus C) Anterior frontal and occipital corpus callosum segments D) Motor corpus callosum segment. E) Left and right Superior Cerebellar Peduncle. Dashed yellow lines represent location of Region of Interest, also indicated with yellow arrows. Colors indicate fiber direction: blue=inferior-superior; green=anterior-posterior, red=left-right.

We selected these tracts *a priori* given that they are easily identifiable with tractography methods in the immature neonatal brain and because of their clinical relevance to preterm birth. Specifically, the CST is a large tract running superiorly to inferiorly, that has been associated with motor development and cerebral palsy in preterm born infants.^31,32^ The ILF is a large transverse tract that has shown associations with language development in preterm infants^33^ and structural alterations in older preterm children.^34,35^ The corpus callosum is the largest white matter structure in the brain. Corpus callosum microstructural alterations are a common finding in the aftermath of preterm birth^3,36^ and have been identified in preterm infants with more inflammation associated conditions.^37,38^ We chose the SCP as representative of cerebellar effects. The SCP is consistently identified in the neonatal preterm population. Cerebellar changes have been identified in preterm infants who received neonatal steroids.^39^

### Statistical Approach

Statistical analyses were conducted using the Statistical Package for the Social Sciences (SPSS version 28.0.1.1, IBM Corp., 2018). Statistical significance was set at p < 0.05. Adherence to the normal distribution of clinical and neurobiological data was assessed using the Shapiro–Wilk test. Based on normality, we used parametric tests for all associations. Descriptive statistics were used to characterize the sample. Chi-square or Fisher’s Exact tests and two-tailed t-tests for independent samples were used to examine differences between Protocol (P) and Non-protocol (NP) on demographic variables. We computed Cohen’s d to assess the effect size of group differences. We analyzed the mean tract metrics FA and MD, averaging left and right tract metrics and planned to use a False Discovery Rate (FDR) of 5% for zero-order associations to account for multiple comparisons.^40^ Power analysis performed in G*Power^41^ for independent two-tailed t-tests confirmed that the final sample size had adequate power (beta =0.8, alpha = 0.05) to detect large effects (Cohen’s d = >.8). Such effect sizes are of comparable magnitude to developmental differences in diffusion metrics found by comparing extremely preterm to very preterm infants at term equivalent age. ^42,43^ In order to rule out the possibility that children who received some steroids in the non-protocol group or who received more or less than the full protocol in the protocol group altered the results, the analyses were repeated using only infants who had no steroids and the full course of steroids. The pattern of results was the same, therefore the results report all children.

## RESULTS

Participant characteristics are shown in **Table 1**. By design, the NP group had significantly lower steroid exposure than the P group (p < 0.001). Participant characteristics were similar in both groups, with no significant differences in gestational age, sex, parent-reported race or ethnicity, antenatal steroid exposure, moderate to severe bronchopulmonary dysplasia^2^, necrotizing enterocolitis or spontaneous intestinal perforation, culture positive sepsis, retinopathy of prematurity (any stage), severe intraventricular hemorrhage (grade 3 or 4), length of stay, or ultrasound findings, or MRI characteristics (all p > 0.05). There was no significant difference between groups in terms of the radiologist’s read MRI findings of no abnormalities versus presence of abnormalities (No abnormalities: NP n=8, P n = 19, χ^2^ p = 0.12). Of the qualitative MRI readings with abnormal findings, most were minor (e.g. ventricular prominence or old blood products in the ventricles. 3 participants in the P group had small foci of blood products in the cerebellum. One participant in the P group had mild to moderate white matter injury and frontoparietal polymicrogyria. One participant in the NP group had unilateral ventriculomegaly.

**Table 1.** Participant Characteristics for the two groups, Non-Protocol, defined as less than 50% of the protocol steroid dose, and Protocol, defined as at least 50% of the protocol dose.

|  | **Non-**<br>**Protocol**<br>**N = 22**<br>**Mean(SD)**<br>**or n (%)** | **Protocol**<br>**N = 33**<br>**Mean(SD)**<br>**or n (%)** | **t(df) or**<br>**$\chi^2$(df)** | **Cohen's**<br>**d** | **p** |
| --- | --- | --- | --- | --- | --- |
| Steroid Exposure (mg/kg) | 0.51 (1.04)<br>min = 0<br>max = 4.00 | 8.42 (1.19)<br>min = 5.00<br>max = 11.00 | -25.3 (53) | -6.95 | <0.001 |
| Birthweight | 864 (162) | 875 (171) | -0.24 (53) | -0.07 | 0.40 |
| Gestational Age, weeks | 26.3 (1.1) | 26.6 (0.8) | -1.30 (53) | -0.36 | 0.20 |
| Female | 9 (41) | 16 (49) | 0.31 (1) |  | 0.58 |
| Race<sup>a</sup> |  |  |  |  | 0.34 |
| Asian/Pacific Islander | 2 (9) | 3 (9) |  |  |  |
| Black or African American | 1 (4) | 0 (0) |  |  |  |
| Other | 13 (59) | 12 (36) |  |  |  |
| Patient Refused/ Unknown | 5 (23) | 14 (42) |  |  |  |
| White | 1 (4) | 4 (12) |  |  |  |
| Ethnicity<sup>b</sup> |  |  |  |  | 0.092 |
| Hispanic/Latino | 11 (50) | 7 (21) |  |  |  |
| Non-Hispanic/Non-Latino | 5 (23) | 13 (39) |  |  |  |
| Patient Refused/Unknown | 6 (26) | 13 (39) |  |  |  |
| Antenatal Steroids<sup>c</sup> | 18 (82) | 32 (97) |  |  | 0.14 |
| Length of Stay, days | 92.1 (21.3) | 84.9 (15.6) | 1.91 (25.9) | 0.61 | 0.067 |
| Discharge Weight, grams | 2960 (768) | 2815 (637) | 1.17 (53) | 0.32 | 0.25 |
| Discharge PMA<sup>d</sup>, weeks | 39.4 (2.6) | 38.8 (2.1) | 1.60 (53) | 0.44 | 0.12 |
| **Medical Complications** |  |  |  |  |  |
| BPD<sup>e</sup> (Moderate/Severe) | 6 (27) | 4 (12) |  |  | 0.18 |
| Necrotizing Enterocolitis | 6 (27) | 7 (21) | 0.27 (1) |  | 0.60 |
| Sepsis | 2 (9) | 5 (15) |  |  | 0.69 |
| Retinopathy of Prematurity | 12 (54) | 18 (54) | 0.00 (1) |  | 1.00 |
| Severe IVH<sup>f</sup> | 3 (14) | 2(6) |  |  | 0.38 |
| Periventricular Leukomalacia | 0 (0) | 0 (0) |  |  |  |
| Cerebellar Hemorrhage | 0 (0) | 0 (0) |  |  |  |
| Seizures | 1 (5) | 0 (0) |  |  | 0.40 |
| **MRI Characteristics** |  |  |  |  |  |
| PMA<sup>c</sup> at Scan, weeks | 37.1 (1.4) | 36.5 (0.8) | 1.92 (30.7) | 0.58 | 0.065 |
| MRI Head Coil |  |  |  |  | 0.074 |
| 8 | 19 (86) | 21 (64) |  |  |  |
| 32 | 3 (14) | 12 (36) |  |  |  |

<sup>a</sup>Parent Reported race, Fisher's Exact Test for Non-White versus White
<sup>b</sup>Parent Reported ethnicity, Fisher's Exact Test for Non-Hispanic/Non-Latino vs Hispanic/Latino
<sup>c</sup>Birth parent received any antenatal steroids, yes/no.
<sup>d</sup>PMA, Post-Menstrual Age
<sup>e</sup>Bronchopulmonary dysplasia as described by Jobe and Bancalari.[2]
<sup>f</sup>Intraventricular Hemorrhage

The proportion of infants with successful tracking was similar between the NP (0.75 ± 0.22) and P (0.84 ± 0.18) groups (t = −1.58, p = 0.12). Mean FA and MD for the average of the left and right tracts and for the anterior, middle, and posterior segments of the corpus callosum are shown in **Table 2**. There were no significant large-effect size differences between the two groups in any metric in any tract. No significant differences between the protocol and non-protocol groups were identified after adjusting for post-menstrual age at scan, and medical complications. We repeated the analyses in the subset of children whose MRI was collected using an 8-channel head coil (n=40). We found no significant differences in mean FA or MD of white matter tracts for the 8-channel head coil subset (data not shown).

**Table 2.** Mean White Matter Tract Microstructural Metrics.

| **White Matter Tract**<br>**Metric** | **Non-Protocol Steroid**<br>**Mean (SD)** | **Protocol Steroid**<br>**Mean (SD)** | **t(df)** | **Cohen's**<br>**d** |
| --- | --- | --- | --- | --- |
| **FA** |  |  |  |  |
| Corticospinal Tract | 0.29 (0.04) (n=18) | 0.29 (0.05) (n=30) | -0.10 (46) | -0.030 |
| Inferior Longitudinal<br>Fasciculus | 0.19 (0.03) (n=21) | 0.19 (0.02) (n=33) | 0.47 (52) | 0.132 |
| Superior Cerebellar<br>Peduncle | 0.20 (0.06 ) (n=21) | 0.18 (0.05) (n=33) | 1.85 (52) | 0.52 |
| Corpus Callosum |  |  |  |  |
| Occipital | 0.29 (0.07) (n=14) | 0.29 (0.07) (n=22) | -0.031 (34) | -0.011 |
| Motor | 0.21 (0.03) (n=14) | 0.21 (0.04) (n=23) | -0.035 (35) | -0.012 |
| Anterior Frontal | 0.22 (0.05) (n=15) | 0.23 (0.05) (n=29) | -0.68 (42) | -0.22 |
| **MD** |  |  |  |  |
| Corticospinal Tract | 1.15 (0.07) (n=18) | 1.16 (0.07) (n=30) | -0.78 (46) | -0.23 |
| Inferior Longitudinal<br>Fasciculus | 1.43 (0.14) (n=21) | 1.43 (0.09) (n=33) | 0.078 (52) | 0.022 |
| Superior Cerebellar<br>Peduncle | 1.16 (0.17) (n=21) | 1.16 (0.16) (n=33) | -0.013 (52) | -0.004 |
| Corpus Callosum |  |  |  |  |
| Occipital | 1.40 (0.07) (n=14) | 1.44 (0.13) (n=22) | -1.08 (34) | -0.37 |
| Motor | 1.43 (0.12) (n=15) | 1.41 (0.10) (n=23) | 0.37 (35) | 0.13 |
| Anterior Frontal | 1.44 (0.10) (n=15) | 1.46 (0.10) (n=29) | -0.65 (42) | -0.21 |

FA, Fractional Anisotropy; MD, Mean Diffusivity

## DISCUSSION

In this retrospective study of 55 extremely preterm born infants who did or did not receive a low-dose early postnatal treatment protocol of prophylactic hydrocortisone for BPD prevention, we report the first assessment of cerebral and cerebellar white matter tract microstructure. Contrary to our initial hypothesis, we identified no significant differences in white matter microstructural metrics at near-term gestational age. Based on our power analysis, we had adequate power to detect a large effect size (Cohen’s d=.8). We cannot conclusively determine if hydrocortisone had a moderate or minimal effect on white matter microstructure in the cerebrum or cerebellum. These results suggest that hydrocortisone, when administered at low dosage during the first 10 days of life, may have minimal impact on white matter maturation at near-term age or were overwhelmed by other factors.

Our findings are interesting in the context of prior studies assessing volumetric brain changes in preterm infants after postnatal hydrocortisone. Unlike studies of dexamethasone administration, most of the studies assessing a larger cumulative early hydrocortisone dose compared with the PREMILOC regimen identify no significant differences in regional brain volumes.^44–47^ There is one exception, by Tam et al,^39^ who identified differences in the change in cerebellar volume from soon after preterm birth until near term age among children who received a similar regimen of hydrocortisone as the PREMILOC protocol. The attenuation in cerebellar growth for infants administered hydrocortisone was less pronounced than that for dexamethasone administration compared to infants who received no steroids. Combined with our findings, it may be that hydrocortisone effects on early brain development are primarily in non-white matter structures.

White matter development is dynamic and rapid in the first two years of life.^48,49^ Clinically relevant white matter differences might be found at a different stage in development. If glucocorticoids decrease neuroinflammation, oligodendrocyte precursor cells may be spared injury and progress through a normal maturational trajectory. Looking beyond infancy into childhood or even young adulthood could reveal differences in ultimate microstructural organization and integrity. On the other hand, steroids themselves can disrupt oligodendrocyte precursor maturation, with higher potency steroids having a more pronounced effect. It is possible that, as was theorized by the PREMILOC study team, low-dose hydrocortisone may not cause alterations to oligodendrocyte precursor cell development in the same way that more potent and high-dose glucocorticoids do.^7,14,50^ This explanation is consistent with recent research looking at brain outcomes related to hydrocortisone treatment. Studies looking at macroscopic brain injuries (PVL, IVH, brain lesions) and brain volume have indicated that hydrocortisone was not associated with adverse effects on brain, in contrast to the reduced brain volume and increased brain abnormalities seen with dexamethasone.^6,7,44^ The protocol instituted in the LPCH NICU was chosen based on clinical consensus, considering the available evidence for preventing bronchopulmonary dysplasia among extremely preterm infants while minimizing potential adverse effects of treatment. There are other early hydrocortisone protocols with higher doses given over longer periods.^51,52^ The cumulative hydrocortisone exposure in these protocols could have detectable effects on brain white matter microstructure. Institutions using higher doses of hydrocortisone should consider examining the effects on brain microstructure.

Several limitations constrain interpretation of our findings. Infants were not randomized to control or treatment groups, but rather received the treatment as a function of timing of birth. There could have been bias in children who were started but did not receive the complete steroid protocol. Six infants in this category had hydrocortisone discontinued or held due to hyperglycemia and three due to hypertension, suggesting that these children may not have had adrenal insufficiency and may have been physiologically more mature than infants who completed the protocol. Neonatal hyperglycemia after preterm birth has been associated with decreased apparent diffusion coefficient in white matter.^53^ While there were no differences in several major neonatal medical conditions in the two groups, this study was not powered to exhaustively account for the many factors that contribute to white matter development. Of 86 eligible infants who received non-protocol or protocol steroids during the study period, 31 (36%) had imaging that was unsuitable for tractography. dMRI processing pipelines are generally developed for mature brains which we adapted to use on infant brain scans. Gray-white matter differentiation is especially underdeveloped in the preterm brain, and likely limited the success of the pipeline. Additionally, the scans used in this study were obtained during clinical care without sedation and primarily intended for radiologist reading. In the face of the constraints of neonatal brain development and clinical imaging, our success in applying advanced, quantitative analytic methods is an important step in moving these methods from the research realm into the clinical sphere. While no significant differences with large effects were found based on hydrocortisone protocol exposure, automated, objective measures of brain structure have potential applications to assessing the effects of other treatments and to outcome prediction in preterm infants.

## CONCLUSION

In this cohort of infants born extremely preterm, a 10-day treatment with early, low-dose hydrocortisone did not relate to differences in early white matter microstructure as indexed by FA and MD. Hydrocortisone treatment may have minimal impact on white matter maturation of the corpus callosum and early neurodevelopmental outcomes. The findings from this research are encouraging for the safety of early, low-dose hydrocortisone as a clinical intervention to potentially reduce the incidence and severity of bronchopulmonary dysplasia.

## Supporting information

Strobe Checklist

## Data Availability

Deidentified data will be made available upon reasonable request made to the corresponding author.

## Author Contributions

Dr. Sarah Dubner conceptualized and designed the study, coordinated and supervised data collection, carried out the data analyses and interpretation, drafted the initial manuscript, and critically reviewed and revised the manuscript.

Lucy Rickerich collected data, performed initial data analyses, drafted the initial manuscript, and critically reviewed and revised the manuscript.

Dawson Sproul collected data and critically reviewed and revised the manuscript.

Rocio Poblaciones performed data analysis and critically reviewed and revised the manuscript.

Dr. Lisa Bruckert supervised and performed data analysis and critically reviewed and revised the manuscript.

Dr. Melissa Scala critically reviewed and revised the manuscript for important intellectual content.

Dr. Heidi M. Feldman conceptualized and designed the study and critically reviewed and revised the manuscript for important intellectual content.

Dr. Katherine E. Travis coordinated and supervised data collection, supervised analysis and critically reviewed and revised the manuscript for important intellectual content.

All authors approved the final manuscript as submitted and agree to be accountable for all aspects of the work.

## Consent Statement

Patient consent was not required for this retrospective study.

## REFERENCES

1. Jobe, A. The new bronchopulmonary dysplasia. Current Opinion in Pediatrics 23, 167–172 (2011).

2. Jobe, A. H. & Bancalari, E. Bronchopulmonary Dysplasia. Am J Respir Crit Care Med 163, 1723–1729 (2001).

3. Volpe, J. J. Brain injury in premature infants: a complex amalgam of destructive and developmental disturbances. Lancet Neurol 8, 110–24 (2009).

4. Watterberg, K. L. & Scott, S. M. Evidence of Early Adrenal Insufficiency in Babies Who Develop Bronchopulmonary Dysplasia. Pediatrics 95, 120–125 (1995).

5. Doyle, L. W. & Anderson, P. J. Long-term outcomes of bronchopulmonary dysplasia. Seminars in Fetal and Neonatal Medicine 14, 391–395 (2009).

6. Baud, O. et al. Effect of early low-dose hydrocortisone on survival without bronchopulmonary dysplasia in extremely preterm infants (PREMILOC): a double-blind, placebo-controlled, multicentre, randomised trial. Lancet 387, 1827–1836 (2016).

7. Alison M et al. Prophylactic hydrocortisone in extremely preterm infants and brain MRI abnormality. Arch Dis Child Fetal Neonatal Ed 105, 520–525 (2020).

8. Baud, O. et al. Association Between Early Low-Dose Hydrocortisone Therapy in Extremely Preterm Neonates and Neurodevelopmental Outcomes at 2 Years of Age. JAMA 317, 1329– 1337 (2017).

9. Baud, O., et al. Two-year neurodevelopmental outcomes of extremely preterm infants treated with early hydrocortisone: treatment effect according to gestational age at birth. Arch. Dis. Child. Fetal Neonatal Ed. 104, F30–F35 (2019).

10. Thompson DK et al. Corpus callosum alterations in very preterm infants: perinatal correlates and 2 year neurodevelopmental outcomes. Neuroimage 59, 3571–81 (2012).

11. Shah, D. K. et al. Adverse neurodevelopment in preterm infants with postnatal sepsis or necrotizing enterocolitis is mediated by white matter abnormalities on magnetic resonance imaging at term. The Journal of pediatrics 153, 170–5, 175.e1 (2008).

12. Volpe, J. J., Kinney, H. C., Jensen, F. E. & Rosenberg, P. A. The developing oligodendrocyte: key cellular target in brain injury in the premature infant. International journal of developmental neuroscience_J: the official journal of the International Society for Developmental Neuroscience 29, 423–40 (2011).

13. Chang, Y. P. Evidence for adverse effect of perinatal glucocorticoid use on the developing brain. Korean J Pediatr 57, 101–109 (2014).

14. Huang, W. L., Dunlop, S. A. & Harper, C. G. Effect of Exogenous Corticosteroids on the Developing Central Nervous System: A Review. Obstetrical & Gynecological Survey 54, 336–342 (1999).

15. Noguchi, K. K. Glucocorticoid Induced Cerebellar Toxicity in the Developing Neonate: Implications for Glucocorticoid Therapy during Bronchopulmonary Dysplasia. Cells 3, 36– 52 (2014).

16. Feldman, H. M., Yeatman, J. D., Lee, E. S., Barde, L. H. & Gaman-Bean, S. Diffusion tensor imaging: a review for pediatric researchers and clinicians. Journal of developmental and behavioral pediatrics_J: JDBP 31, 346–56 (2010).

17. Lerma-Usabiaga, G., Mukherjee, P., Perry, M. L. & Wandell, B. A. Data-science ready, multisite, human diffusion MRI white-matter-tract statistics. Scientific Data 7, 422 (2020).

18. Liu, M., Lerma-Usabiaga, G., Clascá, F. & Paz-Alonso, P. M. Reproducible protocol to obtain and measure first-order relay human thalamic white-matter tracts. Neuroimage 262, 119558 (2022).

19. Zöllei, L., Iglesias, J. E., Ou, Y., Grant, P. E. & Fischl, B. Infant FreeSurfer: An automated segmentation and surface extraction pipeline for T1-weighted neuroimaging data of infants 0-2 years. Neuroimage 218, 116946 (2020).

20. Blesa, M. et al. Parcellation of the Healthy Neonatal Brain into 107 Regions Using Atlas Propagation through Intermediate Time Points in Childhood. Front Neurosci 10, 220 (2016).

21. Veraart, J., Fieremans, E. & Novikov, D. S. Diffusion MRI noise mapping using random matrix theory. Magnetic Resonance in Medicine 76, 1582–1593 (2016).

22. Veraart, J. et al. Denoising of diffusion MRI using random matrix theory. Neuroimage 142, 394–406 (2016).

23. Kellner, E., Dhital, B., Kiselev, V. G. & Reisert, M. Gibbs-ringing artifact removal based on local subvoxel-shifts. Magn Reson Med 76, 1574–1581 (2016).

24. Andersson, J. L. R. & Sotiropoulos, S. N. An integrated approach to correction for off-resonance effects and subject movement in diffusion MR imaging. NeuroImage 125, 1063– 1078 (2016).

25. Yeatman, J. D., Dougherty, R. F., Myall, N. J., Wandell, B. A. & Feldman, H. M. Tract profiles of white matter properties: automating fiber-tract quantification. PloS one 7, e49790 (2012).

26. Tournier, J.-D., Calamante, F. & Connelly, A. Robust determination of the fibre orientation distribution in diffusion MRI: non-negativity constrained super-resolved spherical deconvolution. Neuroimage 35, 1459–1472 (2007).

27. Takemura, H., Caiafa, C. F., Wandell, B. A. & Pestilli, F. Ensemble Tractography. PLOS Computational Biology 12, e1004692 (2016).

28. Tournier, J.-D. et al. MRtrix3: A fast, flexible and open software framework for medical image processing and visualisation. Neuroimage 202, 116137 (2019).

29. Dougherty, R. F. et al. Temporal-callosal pathway diffusivity predicts phonological skills in children. Proceedings of the National Academy of Sciences of the United States of America 104, 8556–61 (2007).

30. Huang, H. et al. DTI tractography based parcellation of white matter: application to the mid-sagittal morphology of corpus callosum. Neuroimage 26, 195–205 (2005).

31. de Kieviet, J. F. et al. A crucial role of altered fractional anisotropy in motor problems of very preterm children. European journal of paediatric neurology_J: EJPN_J: official journal of the European Paediatric Neurology Society 18, 126–33 (2014).

32. Kelly, C. E. et al. Brain structural and microstructural alterations associated with cerebral palsy and motor impairments in adolescents born extremely preterm and/or extremely low birthweight. Dev Med Child Neurol 57, 1168–1175 (2015).

33. Dubner, S. E., Rose, J., Feldman, H. M. & Travis, K. E. Near Term White Matter Microstructure and Language Outcomes in 2-year-old Children Born Preterm. (2019).

34. Feldman, H. M., Lee, E. S., Yeatman, J. D. & Yeom, K. W. Language and reading skills in school-aged children and adolescents born preterm are associated with white matter properties on diffusion tensor imaging. Neuropsychologia 50, 3348–62 (2012).

35. Murner-Lavanchy, I. M. et al. White matter microstructure is associated with language in children born very preterm. Neuroimage Clin 20, 808–822 (2018).

36. Back, S. A. Brain Injury in the Preterm Infant: New Horizons for Pathogenesis and Prevention. Pediatr Neurol 53, 185–92 (2015).

37. Dubner, S. E. et al. White matter microstructure and cognitive outcomes in relation to neonatal inflammation in 6-year-old children born preterm. NeuroImage: Clinical 101832 (2019) doi:10.1016/j.nicl.2019.101832.

38. Shim, S. Y. et al. Altered Microstructure of White Matter Except the Corpus Callosum Is Independent of Prematurity. Neonatology 102, 309–315 (2012).

39. Tam, E. W. Y. et al. Preterm cerebellar growth impairment after postnatal exposure to glucocorticoids. Sci Transl Med 3, 105ra105 (2011).

40. Benjamini, Y. & Hochberg, Y. Controlling the False Discovery Rate: A Practical and Powerful Approach to Multiple Testing. Journal of the Royal Statistical Society. Series B (Methodological) 57, 289–300 (1995).

41. Faul, F., Erdfelder, E., Buchner, A. & Lang, A.-G. Statistical power analyses using G*Power 3.1: Tests for correlation and regression analyses. Behavior Research Methods 41, 1149– 1160 (2009).

42. Dibble, M., Ang, J. Z., Mariga, L., Molloy, E. J. & Bokde, A. L. W. Diffusion Tensor Imaging in Very Preterm, Moderate-Late Preterm and Term-Born Neonates: A Systematic Review. J Pediatr 232, 48–58.e3 (2021).

43. Rose, S. E. et al. Altered white matter diffusion anisotropy in normal and preterm infants at term-equivalent age. Magn Reson Med 60, 761–7 (2008).

44. Benders, M. J. N. L. et al. Brain development of the preterm neonate after neonatal hydrocortisone treatment for chronic lung disease. Pediatr. Res. 66, 555–559 (2009).

45. Kersbergen, K. J. et al. Hydrocortisone treatment for bronchopulmonary dysplasia and brain volumes in preterm infants. J. Pediatr. 163, 666–671.e1 (2013).

46. Parikh, N., Kennedy, K., Lasky, R., McDavid, G. & Tyson, J. Pilot randomized trial of hydrocortisone in ventilator-dependent extremely preterm infants: effects on regional brain volumes. Journal of pediatrics 162, 685_J690.e1 (2013).

47. Rousseau C, Guichard M, Saliba E, Morel B, & Favrais G. Duration of mechanical ventilation is more critical for brain growth than postnatal hydrocortisone in extremely preterm infants. Eur J Pediatr (2021) doi:10.1007/s00431-021-04113-z.

48. Bruckert, L. et al. Age-Dependent White Matter Characteristics of the Cerebellar Peduncles from Infancy Through Adolescence. Cerebellum 18, 372–387 (2019).

49. Yeatman, J. D., Wandell, B. A. & Mezer, A. A. Lifespan maturation and degeneration of human brain white matter. Nat Commun 5, 4932 (2014).

50. Barres, B. A., Lazar, M. A. & Raff, M. C. A novel role for thyroid hormone, glucocorticoids and retinoic acid in timing oligodendrocyte development. Development 120, 1097–1108 (1994).

51. Watterberg, K. L. et al. Hydrocortisone to Improve Survival without Bronchopulmonary Dysplasia. N Engl J Med 386, 1121–1131 (2022).

52. Onland, W. et al. Effect of Hydrocortisone Therapy Initiated 7 to 14 Days After Birth on Mortality or Bronchopulmonary Dysplasia Among Very Preterm Infants Receiving Mechanical Ventilation: a Randomized Clinical Trial. JAMA 321, 354_J363 (2019).

53. Naseh, N. et al. Early Hyperglycemia in Very Preterm Infants Is Associated with Reduced White Matter Volume and Worse Cognitive and Motor Outcomes at 2.5 Years. Neonatology 1–8 (2022) doi:10.1159/000524923.

